# Childhood Adversity and Risk-taking Behaviors in Youth using National Representative Emergency Room Admission and Survey Data

**DOI:** 10.64898/2026.06.24.26356317

**Authors:** Bo Nicolai Lichtenberg, Tjeerd Rudmer De Vries, Claus Thorn Ekstrøm, Naja Hulvej Rod, Jannie Nielsen

## Abstract

**Background:** Childhood adversity can affect propensity to risk-taking behaviors. We aim to investigate the relation between childhood adversity and risk-taking behaviors in youth using emergency room (ER) admissions and survey data.

**Method:** Using the DANLIFE study, we included 1.2 million Danes. Individuals were assigned into five groups based on childhood adversity exposure from ages 0 to 15 years. We applied survival analyses on repeated outcomes to model ER-admissions due to substances, violence and unintentional injury in the full cohort between ages 16 and 24. We applied logistic regression models to weighted survey data on frequent binge drinking, cannabis use, drug use, and unsafe sex in a nested subsample of 34,064 18 year olds from the Danish National Birth Cohort.

**Results:** The high adversity group was at highest risk of ER-admissions due to substances (HR=3.27, 95% CI [3.10, 3.46]), violence (HR=2.67, 95% CI [2.58, 2.76]) and unintentional injuries (HR=1.30, 95% CI [1.28, 1.33]). In the nested subsample, the high adversity was at highest risk of cannabis use (OR=1.59, 95% CI [1.21, 2.09]), drug use (OR=2.44, 95% CI [1.71, 3.49]) and unsafe sex (OR=1.72, 95% CI [1.34, 2.22]), but at lower risk of frequent binge drinking (OR=0.57, 95% CI [0.37, 0.87]).

**Conclusion:** These findings highlight how childhood adversity is associated with increased engagement in and harm from risk-taking behaviors. To prevent inequalities in health in youth, there is a need for interventions and policies that promote child welfare, as well as targeted support for youth with harmful behavioral patterns.

## Introduction

Childhood adversity, such as exposure to material deprivation, death of parent, or parental substance abuse, is a public health issue that has been linked with later life morbidity (1) and early mortality (2). Even in Denmark, a country with an extensive social security system and universal healthcare, it is estimated that 10% of children experience three or more adverse childhood experiences (3).

Risk-taking refers to the propensity to engage in behaviors with potential negative outcomes (4). It has been shown that stress-induced changes in brain architecture caused by exposure to childhood adversity can have permanent effects on important functions such as emotional regulation, learning ability, and executive function (5). These impaired functions can potentially elevate risk-taking, manifesting in risky behaviors such as binge drinking, drug use, violence, and unsafe sex.

Late adolescence and early young adulthood (hereafter referred to as youth) is a developmental period often characterized by increased engagement in risk-taking behaviors (4). During this transitional phase, individuals experience increasing personal and legal autonomy, seek out new boundaries and engage in novel experiences. As part of this, many take risks such as trying substances and engaging in reckless behaviors (4). In youth, these behaviors are often characterized by risk of acute harm, such as injury or poisoning. Not surprisingly, are unintentional injuries, violence, and substance misuse also among leading causes of disability in this age group (6).

Previous research has found that childhood adversity is associated with common risk-taking behaviors in youth (7–13), but limitations remain. To our knowledge, the link between childhood adversity and leading causes of disability in youth associated with risk-taking behaviors, namely unintentional injuries, violence, and substance misuse, has not been properly investigated. Furthermore, previous studies relied on cumulative counts of adverse childhood experiences, an approach that overlooks type, timing and persistence of childhood adversity (14). Lastly, most studies (8–13) are based solely on survey data, where disadvantaged individuals might be underrepresented due to selective non-response (15), and participants might be inclined to provide social desirable responses (16), which can lead to misclassification when dealing with sensitive topics such as childhood experiences and stigmatized and potentially illegal behaviors. To gain better insights into the relationship between childhood adversity and risk-taking behaviors, we need to overcome these limitations.

In this study, we aim to provide a comprehensive picture of the relation between childhood adversity and risk-taking behaviors in youth ages 16 to 24 years, focusing on acute harm from and self-reported engagement in risk-taking behaviors. Acute harm from risk-taking behaviors will be measured as emergency room admissions due to substances, violence, and unintentional injuries in nationwide unselected register-based data on 1.2 million Danes. Engagement in risk-taking behaviors will be measured in a nested sub-sample of the population as self-reported frequent binge drinking, cannabis use, drug use, and unsafe sex. To capture type, timing, and persistence of childhood adversity, we will utilize multidimensional childhood adversity trajectory groups measured over the first 16 years of life.

## METHOD

### Study design, population and data sources

We used data from the register-based DANLIFE cohort study, which comprises information on childhood adversities and background characteristics for individuals born in Denmark between January 1, 1980, and December 31, 2015, totaling 2,223,927 children (3). In this study, we included individuals born between January 1^st^, 1980, and December 31^st^, 2001, to ensure data on entire childhoods (0-16 years). The follow-up period started when a person turned 16 years of age and lasted until the earliest entry of one of the following events: death, emigration, turning 24 years old or end of follow-up (31st of December 2018). After excluding individuals born after December 31, 2001 (n=866,119), individuals who emigrated or died before turning 16 years (n=76,688), and individuals with missing covariates (n=43,275), we had a sample of 1,237,845 individuals (Supplementary Figure S1).

For a sub-sample of the population, data from the 18-years old survey from Danish National Birth Cohort (DNBC-18) were linked to DANLIFE. DNBC-18 includes self-reported data on alcohol intake, cannabis use, drug use and unsafe sex. The background population of the survey were children born in Denmark between 1996 and 2003 (17). Between 2016 and 2021, 89,205 individuals aged 18 years were invited to participate in an online survey and 49,957 responded. In the present study, we only included those born between 1996 and 2001, to align the included birth years of the DNBC and DANLIFE. Hence, after excluding individuals born after 31^st^ of December 2001 (n=10,853), and individuals with missing data (n=5,039), we had a sample of 34,064 individuals (Supplementary Figure S1).

### Childhood adversity

To operationalize childhood adversity, we used childhood adversity trajectory groups previously developed by Rod et al. (2). These were established by applying multi-trajectory modelling to annual counts of childhood adversities across three predefined and expert-informed dimensions: material deprivation (MD), loss or threat of loss, and family dynamics from birth to age 15 (see Supplementary Table S1 for definitions of dimensions and Supplementary Figure S2 for visualization of trajectory groups). Five distinct childhood adversity trajectory groups were identified: *low adversity*, *early life MD*, *persistent MD*, *loss or threat of loss*, and *high adversity* (Figure 1).

**Figure 1.**
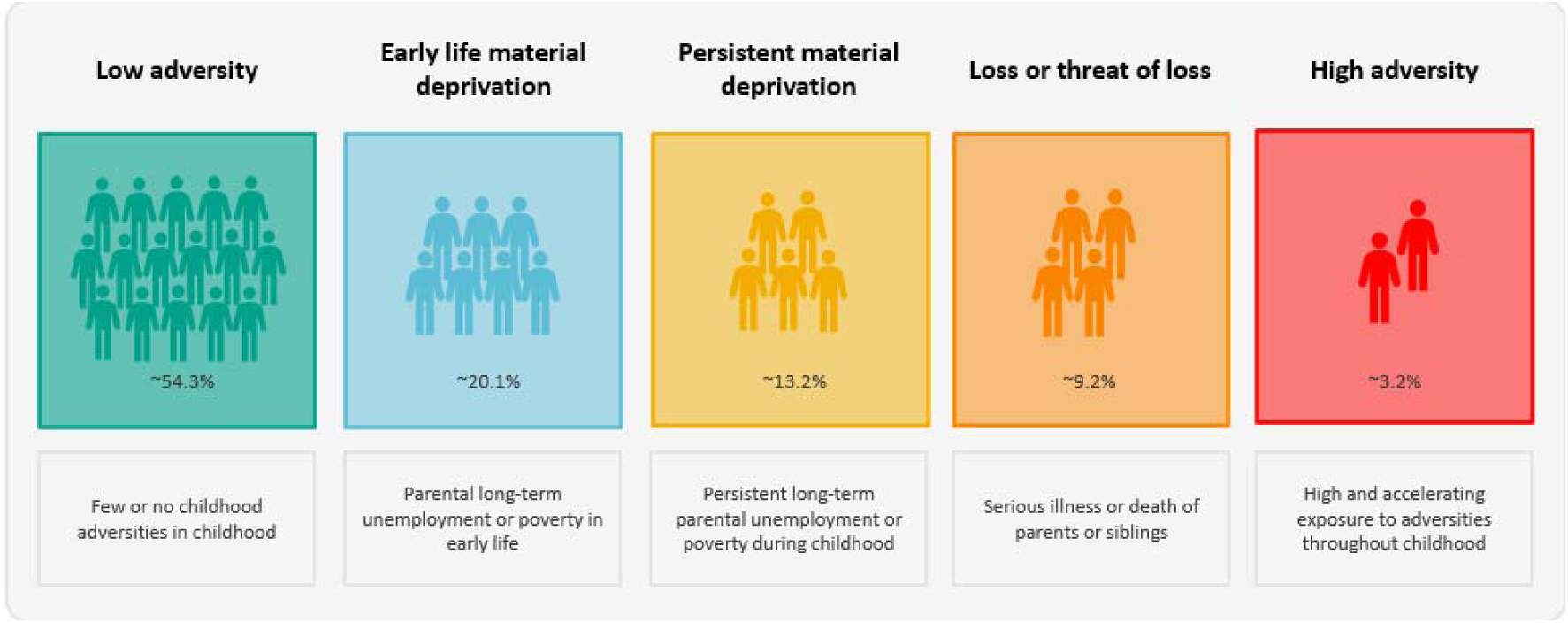
**The five childhood adversity groups and their characteristics.**

### Emergency Room Admissions due to Substances, Violence and Unintentional Injury

Using the Danish National Patient Registry (LPR) (18), we included emergency room (ER) admissions due to substances, violence and unintentional injury.

For each ER admission, we used both primary diagnostic code and secondary diagnostic codes. Because each admission in the LPR can include more than one record, we regard records with discharge and rehospitalization on the same or adjacent day as the same admission, as these likely reflect transfers between hospital departments. For admissions with an external cause of injury, the emergency department uses the Nordic Medico-Statistical Committee’s Classification of External Causes of Injuries, which allows for identification of injuries due to accidents and violence (19).

We measured substance admissions using alcohol and drug-related codes (20,21). We used the contact code for violence to identify ER admissions due to violence (19). We measured unintentional injuries using codes related to injury in ICD-10 chapter S and T and the contact code for accidents (19). See Supplementary Table S2 for detailed outcomes definitions.

### Self-reported frequent binge drinking, cannabis use, drug use, and unsafe sex

From DNBC-18, we included items related to binge drinking, cannabis use, drug use, and unsafe sex. Frequent binge drinking was defined as weekly consumption of 5 or more drinks in one episode. As drinking is normative in Denmark, we opted for this more stringent threshold of weekly binge drinking compared to the commonly used threshold of monthly binge drinking (22). We included items measuring any drug use and cannabis use in the last year. Cannabis was a separate category from the drugs category, as it is a more prevalent drug at this age (23). We included items related to if the individual ever had unprotected sex. This cutoff has been used in a previous study on sexual risk-taking among youth in Denmark (7). All four behaviors were coded as no/yes. See survey questions in Supplementary Table S3.

### Covariates

We included variables expected to confound the association, namely, sex assigned at birth (24), preterm birth (25), household’s highest education at the time of birth (26), origin of parents (predominantly European descent [Europe, North America, Australia, and New Zealand] or non-European descent, if at least one parent had another nationality) (27), and maternal age at time of birth (28).

### Statistical analysis

To assess risk of admissions due to substances, violence, and unintentional injuries, and to account for possibility of repeated admissions, we used the Andersen-Gill time-to-event model for repeated outcomes (29) to obtain hazard ratios (HRs) and 95% confidence intervals. The Andersen-Gill method allows the same individual to have multiple contacts during the follow-up period. We used robust standard errors (variance-corrected method) to account for intra-individual correlation. We checked the proportional hazard assumption using log-log plots, which was deemed fulfilled. We additionally used the semi-parametric additive hazard model by McKeague and Sasieni (30) to obtain hazard differences (HDs) and 95% confidence intervals. Age 16 to 24 was the underlying time scale for both models. We excluded risk-time while individuals were under any hospitalization. We censored at death, emigration, turning 24 years old or end of follow-up 31st of December 2018.

We used multivariable logistic regression with a quasibinomial link to obtain odds-ratio (OR) and 95% confidence intervals for frequent binge drinking, cannabis use, drug use, and unsafe sex. To show adjusted percentage points differences (PPD) and 95% confidence intervals by adversity group for each outcome, we estimated average marginal effects (31). To reduce potential selection bias, the sample was weighed using inverse probability weighting derived from a logistic regression according to the background population (312,502) individuals born between 1996-2001. The procedure and model checks are outlined in Supplementary Material S4.

The low adversity group was the reference group in all models. All models were adjusted for the covariates, except for parental origin in the DNBC-18 sample, as there were no parents born outside Europe in the high adversity group. No adjustment for multiple testing was done.

### Sensitivity analyses

We first repeated the analyses without adjusting for parental education, as parental education is highly correlated with material deprivation. Second, to examine potential selection bias and effectiveness of the weights, we investigated if violence and unintentional injury admissions differed by weighted, unweighted, and background population in incidence rate ratios. Due to low case counts we did not include substance admissions in this analysis as it makes it difficult to rule out random error in the estimates. Lastly, we stratified the analyses by sex to test if the results were driven by either men or women.

All analyses were done in Stata 18 and R (4.5.0) using packages timereg (2.0.6), survey (4.4.2), and marginaleffects (0.26.0).

### Ethical approval

The DANLIFE study received approval from the Danish Data Protection Agency via the joint notification submitted by the Faculty of Health and Medical Sciences, University of Copenhagen (record no. 514-0641/21-3000).

The Danish National Birth Cohort (DNBC) cohort is approved by the Danish Data Protection Agency under the general approval (fællesfortegnelse) granted to Statens Serum Institut, reference number 18/04608 and by the Committee on Health Research Ethics (case number (KF) 01-471/94).

## RESULTS

### Descriptive statistics

Descriptive statistics by adversity groups are shown in table 1. A higher proportion in the high adversity group came from households with low parental education, had younger mothers, and were more frequently born preterm. The low adversity group, and to a lesser extent the loss or threat of loss group, were characterized by older maternal age and higher parental education. These patterns were similar for the register-based population and the DNBC-18 population.

**Table 1.** Descriptive characteristics of outcomes and covariates across childhood adversity trajectory groups in the register population (N=1,237,845) and DNBC-18 population (N=34,064).

|  |  | Low adversity | Early life MD | Persistent MD | Loss or threat of loss | High adversity |
| --- | --- | --- | --- | --- | --- | --- |
| **DANLIFE register population** |  |  |  |  |  |  |
| **Individuals** | n | 671,967 | 248,934 | 163,501 | 113,866 | 39,577 |
|  | (%) | (54.3) | (20.1) | (13.2) | (9.2) | (3.2) |
|  | Person years | 4,336,747 | 1,730,229 | 1,158,890 | 782,727 | 272,204 |
| **ER outcomes** |  |  |  |  |  |  |
| Substance | Individuals admitted ≥1 (%) | 1.7 | 2.6 | 3.1 | 3.1 | 6.5 |
| Violence | Individuals admitted ≥1 (%) | 4.1 | 6.3 | 8.4 | 6.9 | 13.7 |
| Unintentional Injury | Individuals admitted ≥1 (%) | 44.5 | 49.5 | 51.9 | 48.8 | 57.8 |
| **Covariates** |  |  |  |  |  |  |
| Sex | Male (%) | 51.3 | 51.1 | 51.1 | 51.0 | 54.2 |
| Highest household education | Low (%) | 8.0 | 21.4 | 32.4 | 21.0 | 53.8 |
|  | Medium (%) | 48.1 | 54.9 | 48.6 | 50.1 | 35.6 |
|  | High (%) | 43.6 | 23.4 | 18.3 | 28.5 | 9.7 |
| Parental origin | Non-western (%) | 1.1 | 3.5 | 8.4 | 3.5 | 2.0 |
| Maternal age | <20 years old (%) | ≤1 | 3.5 | 6.8 | 3.4 | 10.5 |
|  | 20-30 years old (%) | 64.8 | 72.9 | 70.8 | 63.3 | 67.7 |
|  | >30 years old (%) | 34.2 | 23.6 | 22.3 | 33.3 | 21.8 |
| Preterm | <37 weeks (%) | 4.8 | 5.3 | 5.5 | 7.0 | 8.7 |
| **DNBC-18 survey population (Weighted)** |  |  |  |  |  |  |
| **Individuals** | N | 24180 | 4322 | 1652 | 3386 | 523 |
|  | (%) | (71.0%) | (12.7%) | (4.8%) | (9.9%) | (1.5%) |
| **Self-reported outcomes** |  |  |  |  |  |  |
| Frequent binge drinking | Weekly binge drinking (%) | 16.7 | 14.7 | 14.1 | 14.5 | 10.2 |
| Cannabis Use | Any cannabis use in the last year (%) | 23.6 | 27.3 | 27.0 | 30.7 | 33.0 |
| Drug use | Any drug use in the last year (%) | 8.0 | 9.3 | 9.8 | 13.0 | 17.7 |
| Unsafe sex | Unprotected sex one or more times (%) | 40.9 | 45.1 | 45.4 | 49.9 | 55.9 |
| **Covariates** |  |  |  |  |  |  |
| Sex | Male (%) | 51.1 | 50.2 | 48.9 | 47.9 | 52.4 |
| Highest household education | Low (%) | 6.3 | 21.9 | 21.7 | 20.1 | 52.7 |
|  | Medium (%) | 47.8 | 51.8 | 55.7 | 51.0 | 38.2 |
|  | High (%) | 45.8 | 26.3 | 22.6 | 28.9 | 9.1 |
| Parental origin<sup>a</sup> | Non-western (%) | - | - | - | - | - |
| Maternal age | <20 years old (%) | ≤1 | 3.3 | 3.3 | 2.9 | 9.4 |
|  | 20-30 years old (%) | 58.3 | 61.2 | 60.5 | 56.9 | 65.7 |
|  | >30 years old (%) | 41.2 | 35.5 | 36.2 | 40.2 | 24.9 |
| Preterm | <37 weeks (%) | 4.7 | 5.6 | 5.8 | 8.7 | 14.1 |

Note. a=Excluded due to lack of observations in the high adversity group.

Admission proportions differed by adversity groups; the high adversity group had the highest proportions of one or more admissions due to substance (6.5%), violence (13.7%), and unintentional injuries (57.8%), while the low adversity group had the lowest proportions for substances (1.7%), violence (4.1%), and unintentional injuries (44.5%). The DNBC-18 population showed similar patterns. The high adversity group reported the highest proportion of cannabis use (33.0%), drug use (17.7%) and unsafe sex (55.9%), while the low adversity group had the lowest proportion for cannabis use (23.6%), drug use (8.0%) and unsafe sex (40.9%). Conversely, the low adversity group had the highest proportion reporting frequent binge-drinking (16.7%), while the high adversity group had the lowest proportion (10.2%).

### Substance, violence, and unintentional injury emergency room admissions

Figure 2 presents age-specific rates for substance use, violence, and unintentional injury admissions. Across all adversity groups, rates of substance use and violence admissions peaked around ages 18–19 and then steadily declined, whereas rates of unintentional injury admissions declined steadily from ages 16 to 24. The incidence rates and especially the peak are most pronounced for the high adversity group.

**Figure 2.**
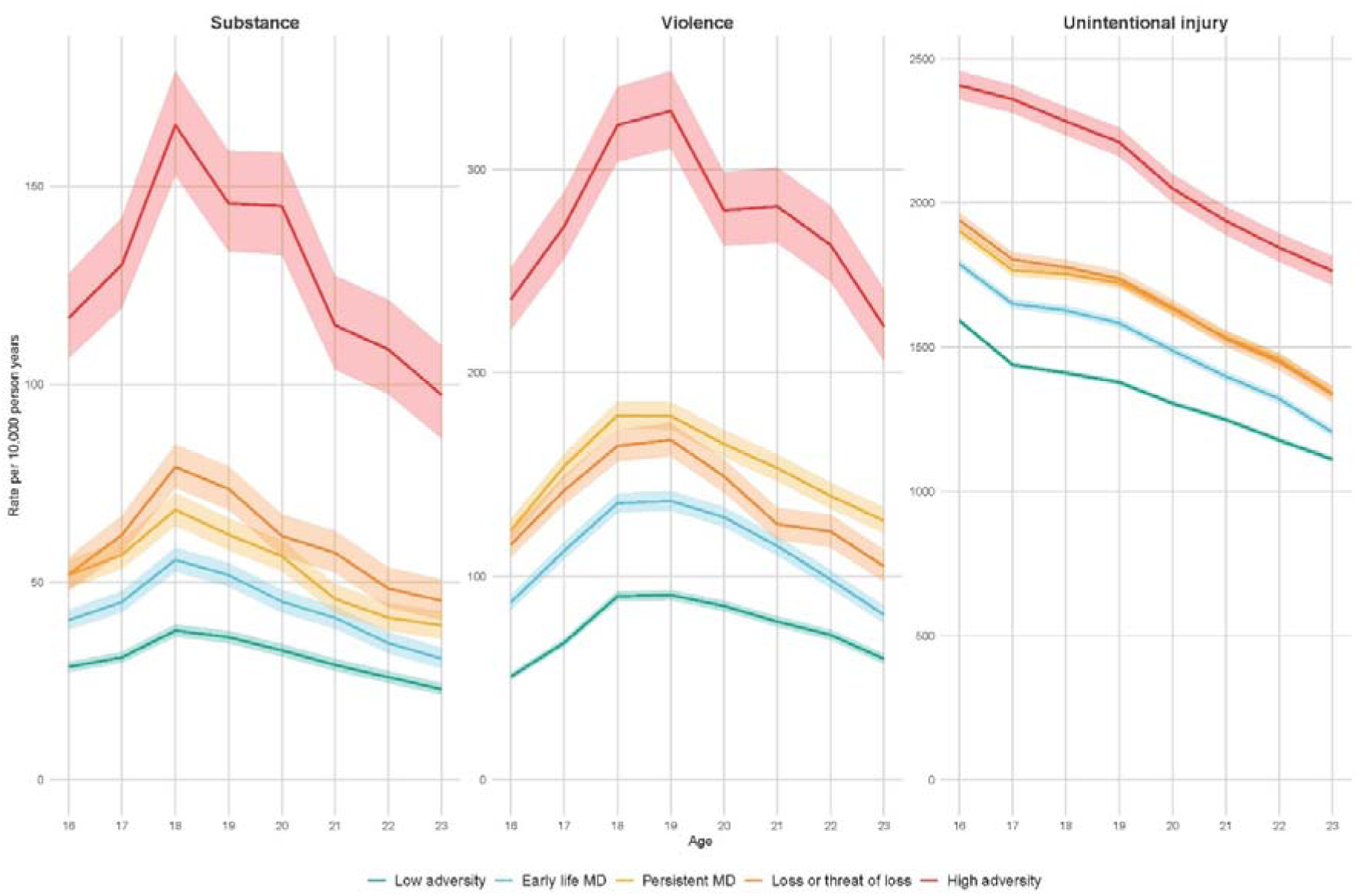
Age specific incidence rates per 10,000 individuals for substance, violence, and unintentional injury ER-admissions.

With the low adversity group as reference, the high adversity group showed the highest risks across all ER outcomes (Figure 3) with a HR of 3.27 (95% CI [3.10, 3.46]) for substance admissions, 2.67 (95% CI, [2.58, 2.76]) for violence admissions, and 1.30 (95% CI, [1.28, 1.33]) for unintentional injury admissions, corresponding to 86 (95% CI [82, 91]), 162 (95% CI [156, 168]) and 502 (95% CI [484, 520]) additional cases per 10,000 person years, respectively. We also observed that the loss or threat of loss group had the second highest risk of substance admissions (HR=1.82, 95% CI [1.16, 1.19]; HD per 10,000=26, 95% CI [25, 28]) and unintentional injury admissions (HR=1.17, 95% CI [1.64, 1.72]; HD per 10,000=242, 95% CI [232, 252]) followed by the persistent MD group (substance: HR=1.51, 95% CI [1.46, 1.57]; HD per 10,000=16, 95% CI [15, 18]. Unintentional injury: HR=1.10, 95% CI [1.09, 1.11]; HD per 10,000=146, 95% CI [137, 155]) and early life MD group (substance: HR=1.29, 95% CI [1.25, 1.34]; HD per 10,000=9, 95% CI [8, 10]. Unintentional injury: HR=1.04, 95% CI [1.04, 1.05]; HD per 10,000=58, 95% CI [52, 65]). For violence admissions, the loss or threat of loss group (HR=1.68, 95% CI [1.64, 1.73]; HD per 10,000=53, 95% CI [50, 56]) and persistent MD group (HR=1.68, 95% CI [1.64, 1.72]; HD per 10,000= 55, 95% CI [53, 58]) had similar higher risk, and with early life MD showing more modest associations (HR=1.33, 95% CI [1.30, 1.36]; HD per 10,000=24, 95% CI [22, 26]).

**Figure 3.**
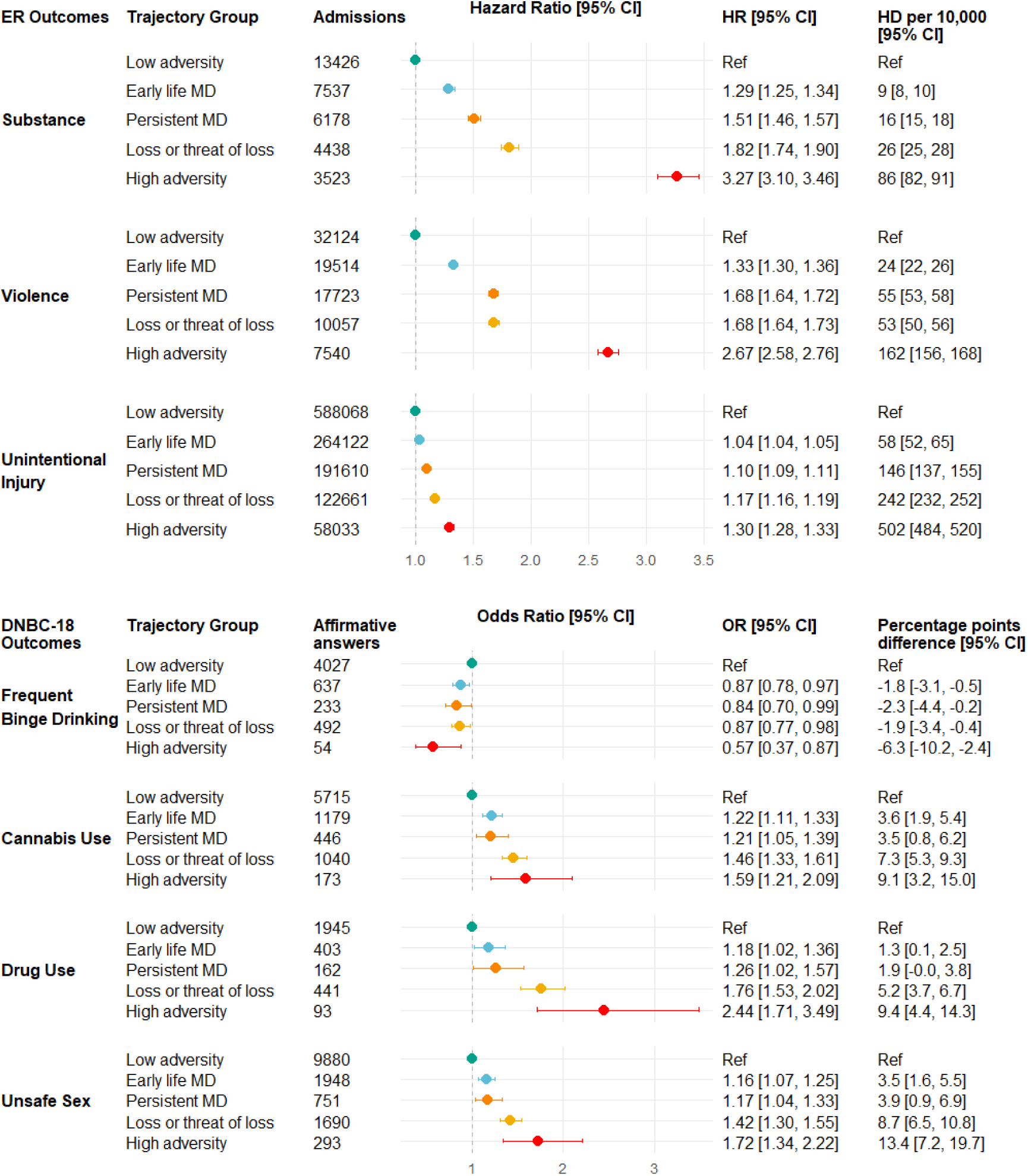
HR and HD per 10,000 individuals for substance, violence, and unintentional injury ER-admissions. ORs and percentage points difference (PPD) for frequent binge drinking, cannabis use, drug use, and unsafe sex. Note. Substance, violence, and unintentional injuries are adjusted for sex, highest household education, parental origin, maternal age at birth, and preterm birth. See Supplementary table S5 for results of alcohol and drug admissions separately. Frequent binge drinking, cannabis use, drug use, and unsafe sex are adjusted for sex, highest household education, maternal age at birth, and preterm birth.

### Frequent binge drinking, cannabis use, drug use, and unsafe sex

In Figure 3, we observed statistically significant differences for frequent binge drinking across childhood adversity groups, where the high adversity group showed the lowest risk (OR=0.57, 95% CI [0.37, 0.87]; PPD= 6.3, 95% CI [-10.2, -2.4]). In contrast, childhood adversity was associated with higher odds of cannabis use, drug use and unsafe sex. The high adversity group had the highest odds and percentage points difference for cannabis use (OR=1.59, 95% CI [1.21, 2.09]; PPD=9.1, 95% CI [3.2,15.0], drug use (OR=2.44, 95% CI [1.71, 3.49]; PPD=9.4, 95% CI [4.4, 14.3]), and unsafe sex (OR=1.72, 95% CI [1.34, 2.22]; PPD=13.4, 95% CI [7.2, 19.7]), followed by the loss or threat of loss group (cannabis use: OR=1.46, 95% CI [1.33, 1.61]; PPD=7.3, 95% CI [5.3, 9.3]. Drug use: OR=1.76, 95% CI [1.53, 2.02]; PPD=5.2, 95% CI [3.7, 6.7]. Unsafe sex: OR=1.42, 95% CI [1.30, 1.55]; PPD=8.7, 95% CI [6.5, 10.8]). The persistent MD and early life MD groups showed more modest but still elevated risk of cannabis use, drug use and unsafe sex.

### Sensitivity analysis

Not adjusting for parental education resulted in slightly stronger associations, except for frequent binge-drinking and cannabis use, which remained similar (Supplementary Table S6A-B). The weighted sample mostly showed greater similarity to the background population than the unweighted estimates. However, for violence admissions, the weighted estimates showed a larger negative deviation from the background population than the unweighted estimates in the high adversity group and the early life MD group (Supplementary Table S7).

*S*tratifying by sex showed similar differences on the additive scale between men and women for substance, violence, and unintentional injury admissions, as well as for cannabis use, and drug use. Differences on the relative scale was driven by lower case count among women. For unsafe sex, childhood adversity was associated with a higher risk among women than men. For example, women in the high adversity group had an OR of 1.96 (95% CI [1.39, 2.76]) and percentage points difference of 16.5 (95% CI [8.3, 24.8]), whereas males had an OR of 1.22 (95% CI [4.9, 14.2]) and a percentage points difference of 4.9 (95% CI [-4.4, 14.2]). For frequent binge drinking, the statistically significant association was mainly driven by men in the high adversity group. See supplementary Table S8A-E for stratification of all outcomes.

## DISCUSSION

This study investigated the association between childhood adversity and risk-taking behaviors in youth by combining registry and survey data. We found that childhood adversity is generally associated with higher risk of acute harm and self-reported engagement in risk-taking behaviors. Especially, the high adversity group, with cumulative and accelerating adversity, showed the highest risk of emergency room admissions due to substances, violence, and unintentional injuries as well as the highest risk of self-reported engagement in cannabis use, drug use and unsafe sex. The latter primarily among women. Contrary to our hypothesis, high adversity showed a lower risk of frequent binge drinking driven by men in the sample.

Our findings align with previous smaller studies on childhood adversity and risk-taking in youth. Andreasen et al. (2024) found that men and women aged 15–29 exposed to ≥3 childhood adversities had higher odds of engaging in unsafe sex (7). Previous studies also found higher risk of drug use by cumulative counts of adverse childhood experiences (8,13). The lower risk of frequent binge drinking for the high adversity group does not align with previous studies (10,11). We add to this body of work by investigating acute harm from and self-reported engagement in risk-taking behaviors, as well as employing multidimensional trajectories of childhood adversity.

The results highlight how childhood adversity is associated with heightened engagement in risk-taking behaviors, which may contribute to inequalities in health in youth. The noticeable higher risk in the high adversity group is likely driven by a myriad of problems that accumulate and interact in childhood and continue through youth. For example, growing up in a deprived environment can cause preference for immediate rewards over long-term benefits (32), increasing likelihood of risk-taking behaviors for short-term purposes, such as relief of stress. Furthermore, witnessing or experiencing violence in childhood, might lead to behavioral modelling and maladaptive beliefs about violence (33,34), which may explain the higher risk of violence admissions in the high adversity group. Childhood adversity can also lead to continued marginalization in youth, predisposing some to risk-taking. For example, a previous study found that the high adversity group had higher risk of leaving school early and long-term dependence on social benefits (35). The stress from such social marginalization and precarity can limit cognitive resources to avoid harmful behaviors (36).

Our findings point to a distinction between socially normative forms of risk-taking and more marginalized and harmful risk behaviors, with childhood adversity primarily associated with the latter. For example, in Denmark alcohol is legal for youth from age 16 years, and binge drinking is a socially accepted and a peer-driven activity (37). Therefore, not participating in binge drinking at this age could be an indication of social isolation rather than an aversion to risk-taking, potentially explaining the diverting pattern of lower odds of frequent binge drinking in the high adversity group. This may also account for the differing findings between this study and previous studies (10,11), which were conducted in the US, where alcohol is not legal before age 21 years. Lastly, the higher risk of substance admissions by childhood adversity suggests more harmful use of substances. Combined, this paints a broader picture of vulnerable groups of youth with complex patterns of harmful behaviors.

The association between childhood adversity and unsafe sex was mainly seen in women. This may reflect gendered power dynamics that tend to give men greater sexual self-determination and freedom than women, for example more agency to negotiate contraceptive use (38). As such, when gender intersects with childhood adversity, it can reinforce power imbalances, leaving women from highly adverse backgrounds vulnerable in negotiating contraceptive use.

Our study has several strengths. By combining emergency room admissions with self-report data, we captured both acute harm from risk-taking behaviors and self-reported engagement in risk-taking behaviors. Furthermore, using registry data allowed us to capture type, timing, and persistence of childhood adversity exposure, revealing a highly vulnerable group characterized by accumulating and accelerating childhood adversity that faced noticeable higher risks across nearly all outcomes. Finally, our use of population-level data enabled us to investigate the association between childhood adversity and leading causes of disability associated with risk-taking in youth, including unintentional injuries, violence and substance use, in an unselected sample.

This study also has limitations. Although using registry data is beneficial for capturing unselected population-level patterns, it can lack specificity. For example, attributing ER-admissions to specific risk-taking behaviors is difficult, as admissions can result from clusters of behaviors, as well as causes beyond individual risk-taking, such as exposure to random violence. Furthermore, by restricting to individuals born in Denmark, we excluded groups such as refugees and immigrants. These groups may face higher risk of MD or loss or threat of loss, and may hold different cultural norms regarding risk-taking behaviors, potentially altering associations. Finally, non-response bias could affect the results in DNBC-18, as individuals with more adverse background might select out of the survey, potentially underestimating the relationship. We addressed this by applying sample weights.

In conclusion, combining register and self-reported data shows that childhood adversity is linked to heightened harm from and engagement in risk-taking behaviors, particularly more harmful and marginalized forms rather than socially normative ones. These findings highlight the need for early interventions that promote child welfare and targeted support for youth with harmful behavioral patterns. The findings also emphasize the importance of cross-sectoral policies addressing the multidimensional nature of childhood adversity to prevent avoidable morbidity and reduce health inequalities.

## Data Availability

The data material contains sensitive and personally identifiable information, why it cannot be made publicly available. Inquiries about secure access to the DANLIFE data under the stated conditions by the Danish Data Protection Agency can be directed at Naja Hulvej Rod.

