## Supplementary Material for "Childhood Adversity and Risk-taking Behaviors in Youth using National Representative Emergency Room Admission and Survey Data"

**^1^ Copenhagen Health Complexity Center, Department of Public Health, University of Copenhagen, Denmark**

**^2^ Section of Biostatistics, Department of Public Health, University of Copenhagen, Denmark**

**^3^ Section for Social Medicine, Department of Public Health, University of Copenhagen, Denmark**

### **Supplementary Figure 1. Flowcharts of included individuals in the full sample and nested sub-sample.**

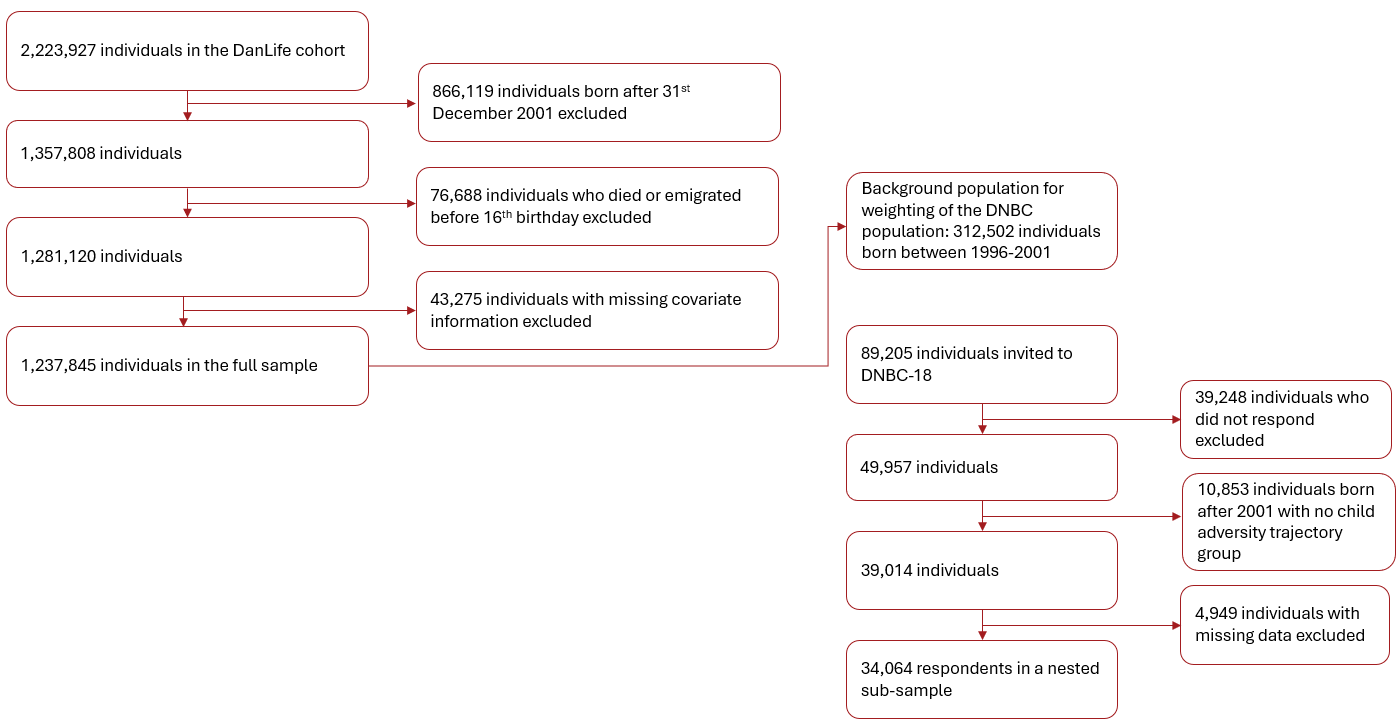

### **Supplementary Table 1. Dimensions and definitions of childhood adversities in the five trajectory group.**

Dimensions and definitions of childhood adversities in the five trajectory groups of adversity identified by Rod et al. (1), with details on the data described in Bengtsson et al. (2). Rod et al. included experts in stress, child health, and child psychology and this panel of experts decided on the three predefined dimensions of childhood adversity.

| Dimension | Adversity | Definition | Registers |
| --- | --- | --- | --- |
| Material deprivation | Family poverty | Family income below 50% of the median national family income in a given year | The Income Statistics Register (3) |
|  | Parental long-term unemployment | A parent being unemployed for at least 12 months | The Integrated Database for Labour Market Research (4) |
| Loss or threat of loss | Death of parent | Death of a parent | The Danish Civil Registration System (5) |
|  | Death of sibling | Death of a sibling | The Danish Civil Registration System (5) |
|  | Parental somatic illness | A parent being diagnosed with one of the diseases included in the Charlson comorbidity index | The Danish National Patient Register (6) |
|  | Parental somatic illness | A sibling being diagnosed with one of the seven  somatic illnesses most commonly related to mortality  in children aged 0–18 years in Denmark:  malignant neoplasm; congenital anomalies of the  heart and circulatory system; congenital anomalies of  the nervous system; cerebral palsy; epilepsy;  cardiomyopathy; congenital disorders of lipid  metabolism | The Danish National Patient Register (6) |
| Family dynamics | Foster care | Being placed in out-of-home care | The Register of Support for Children and Adolescents (7) |
|  | Parental psychiatric illness | A parent being admitted for at least 1 day to a psychiatric hospital or ward with a primary diagnosis related to psychiatric illness (excluding primary diagnoses related to alcohol and drug abuse) | The Danish Psychiatric Central Research Register (8); The Danish National Patient Register (6) |
|  | Sibling psychiatric illness | A sibling being admitted for at least 1 day to a psychiatric hospital or ward with a primary diagnosis related to psychiatric illness | The Danish Psychiatric Central Research Register (8); The Danish National Patient Register (6) |
|  | Parental alcohol abuse | A parent being diagnosed with a disease related to alcohol abuse or buying a prescribed drug used in treatment of alcohol dependence | The Danish Psychiatric Central Research Register (8); The Danish National Patient Register (6); The Danish National Prescription Registry (9) |
|  | Parental drug abuse | A parent being diagnosed with a disease related to drug abuse or buying a prescribed drug used in treatment of drug dependence | The Danish Psychiatric Central Research Register (8); The Danish National Patient Register (6); The Danish National Prescription Registry (9) |
|  | Maternal separation | The mother no longer sharing address with a partner | The Danish Civil Registration System (5) |

Note: This table is from the supplementary material of Rod NH, Bengtsson J, Elsenburg LK, Taylor-Robinson D, Rieckmann A. Hospitalisation patterns among children exposed to childhood adversity: a population-based cohort study of half a million children. Lancet Public Health 2021; published online Sept 29. http://dx.doi.org/10.1016/S2468-2667(21)00158-4.

**Supplementary Figure 2. Childhood adversity trajectories groups.**

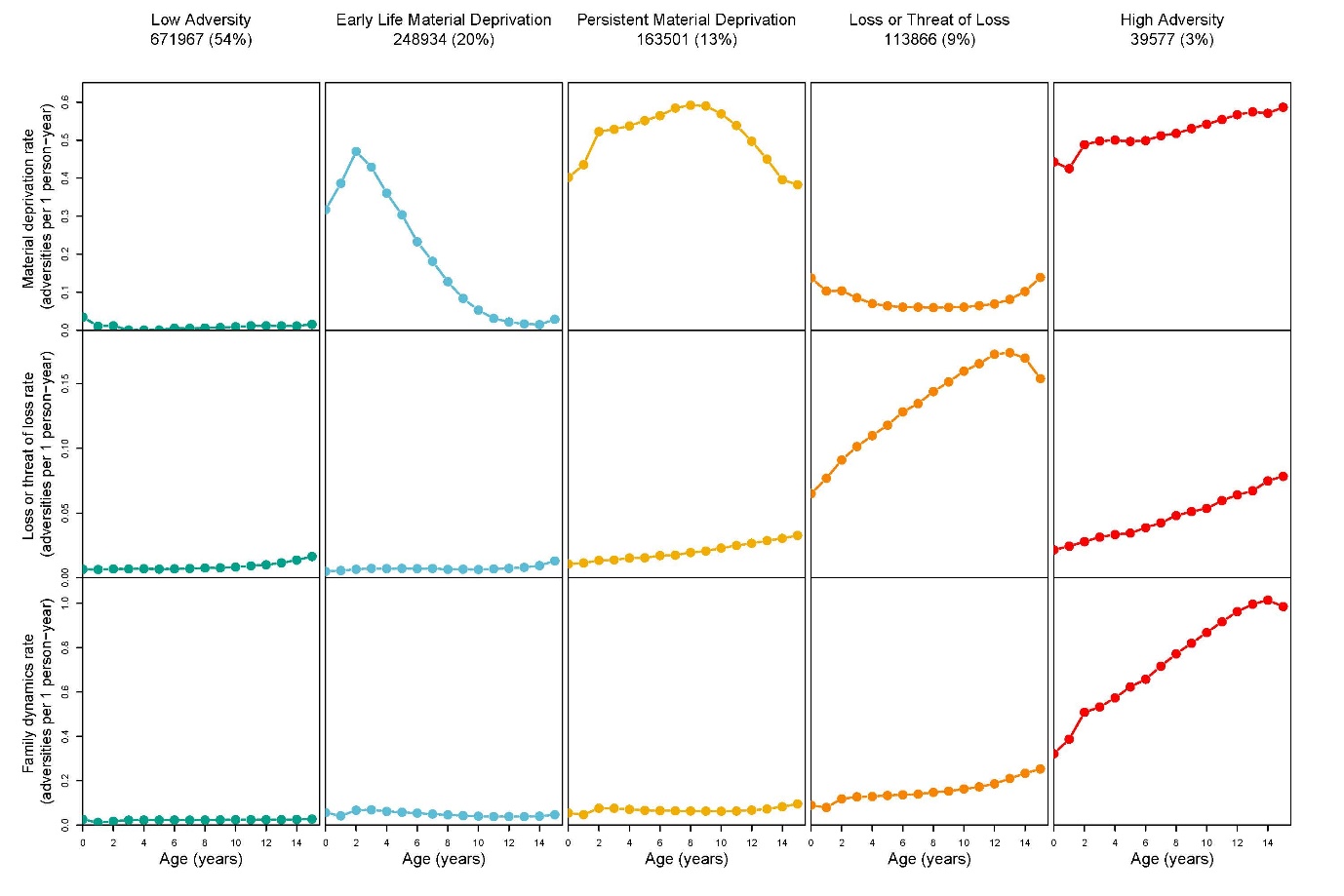

Note. The rate across each childhood adversity dimension for the five childhood adversity trajectory groups.

### **Supplementary Table 2. Substance, violence, and unintentional injury admissions.**

| Outcome | ICD-codes | Reason for admission |
| --- | --- | --- |
| Substance-related admission(1,2) | - F10–F16, F18, F19 - T40-T43, T50, T51 - X40-45 - Y15 - R78.0 | NA |
| Violence-related admission | NA | Violence |
| Unintentional injury-related admission(3) | - DS00–DT98   Excl. injuries due to medical treatment or toxic effects of substances:   - T36–T51 - T79 - T80–T89 - T90–T98 | Accident |

1.	Maharaj T, Fitzgerald N, Gilligan E, Quirke M, MacHale S, Ryan JD. Alcohol-related emergency department presentations and hospital admissions around the time of minimum unit pricing in Ireland. Public Health. 2024 Feb 1;227:38–41.

2.	Wood DM, De La Rue L, Hosin AA, Jurgens G, Liakoni E, Heyerdahl F, et al. Poor Identification of Emergency Department Acute Recreational Drug Toxicity Presentations Using Routine Hospital Coding Systems: the Experience in Denmark, Switzerland and the UK. J Med Toxicol Off J Am Coll Med Toxicol. 2019 Apr;15(2):112–20. doi:10.1007/s13181-018-0687-z

3.	Kruckow S, Santini ZI, Hjarnaa L, Becker U, Andersen O, Tolstrup JS. Associations between alcohol intake and hospital contacts due to alcohol and unintentional injuries in 71,025 Danish adolescents – a prospective cohort study. eClinicalMedicine;64. Available from: https://www.thelancet.com/journals/eclinm/article/PIIS2589-5370(23)00364-4/fulltext

### **Supplementary Table 3. DNBC-18 survey questions.**

| Outcome | Question | Cutoffs |
| --- | --- | --- |
| Frequent binge drinking | Based on the question ‘How often do you drink 5 alcoholic drinks or more at the same occasion?’  Response options: ‘Never’, ‘Less often than once a month’, ‘Monthly’, ‘Weekly’, ‘Daily or almost daily’, ‘Not applicable’ | Affirmative answer weekly or more often. |
| Cannabis use | Based on the question ‘How many times have you used weed within the last year’  Response options included were ‘Not within the last year’, ‘Once, ‘Less than once a month’, ‘Monthly, ‘Weekly’, ‘Daily or almost daily’, and ‘Not applicable’ | Affirmative answer for once or more often. |
| Drug use | Based on the question ‘Have you tried any of the following drugs within the past year’.  Response options included were ‘amphetamine', ‘ecstasy/MDA/, fantasy or similar’, ‘coke’, ‘LSD’, ‘opioids’, mushrooms’, and ‘inhaling of nitrous oxide or solvents’ | Affirmative answer for any of the drugs within the last year. |
| Unsafe sex | Based on the question ‘How many times have you had unprotected sex (not used any kind of protection)?’  Response options: ‘Zero times’, ‘A single time’, ‘2-5 times’, ‘6-10 times’, ‘More than 10 times’, ‘Do not know’, ‘Not applicable’ | Affirmative answer a single or more times. |

### **Supplementary Material 4. Weighting procedure.**

The background population were individuals born from the general population between 1996 and 2001 (N = 364,028). We estimated probability of participation based on highest parental education, maternal age at birth, preterm birth, sex, and any childhood or adolescent psychiatric diagnosis before the age of 18 (Table 1). Weights were rescaled to a mean of 1 and truncated at the 99^th^ percentile. The standardized mean difference (SMD) was calculated to evaluate the balance of covariates between the weighted and unweighted samples. An SMD of less than 0.1 indicated an acceptable covariate balance (1).

Supplementary Table 4A: Predictors included for inverse probability weighting across the unweighted (U) and weighted (W) DNBC-18 population, background (B).

|  | Low adversity | | | Early life MD | | | Persistent MD | | | Loss or threat of loss | | | High adversity | | | SMD |
| --- | --- | --- | --- | --- | --- | --- | --- | --- | --- | --- | --- | --- | --- | --- | --- | --- |
|  | U^a^ | W^b^ | B^c^ | U^a^ | W^b^ | B^c^ | U^a^ | W^b^ | B^c^ | U^a^ | W^b^ | B^c^ | U^a^ | W^b^ | B^c^ |  |
| Individuals (%) | 25531 (75.0) | 24180 (71.0) | 195961 (62.7) | 3763 (11.1) | 4322  (12.7) | 55549 (15.3) | 1416 (4.2) | 1652  (4.8) | 25235 (8.1) | 3034 (8.9) | 3386  (9.9) | 34988 (11.2) | 320 (>1) | 523 (1.5) | 8481 (2.7) |  |
| Sex |  |  |  |  |  |  |  |  |  |  |  |  |  |  |  |  |
| Male | 58.0 | 48.9 | 50.9 | 58.7 | 50.2 | 50.9 | 60.0 | 47.9 | 48.9 | 60.8 | 51.7 | 50.6 | 56.2 | 52.7 | 53.7 | 0,02 |
| Maternal age at birth |  |  |  |  |  |  |  |  |  |  |  |  |  |  |  |  |
| <20 | ≤1 | ≤1 | ≤1 | 1.3 | 3.3 | 2.9 | 1.2 | 3.3 | 3.9 | 1.2 | 2.9 | 2.8 | 4.7 | 9.4 | 8.5 | -0,00 |
| 20-30 | 55.0 | 58.3 | 57.8 | 57.9 | 61.2 | 63.0 | 56.5 | 60.5 | 63.0 | 53.3 | 56.9 | 57.9 | 60.0 | 65.7 | 62.6 | -0,01 |
| >30 | 44.8 | 41.2 | 41.6 | 40.8 | 35.5 | 34.2 | 42.3 | 36.2 | 33.2 | 45.6 | 40.2 | 39.3 | 35.3 | 24.9 | 29.0 | 0,01 |
| Highest household education |  |  |  |  |  |  |  |  |  |  |  |  |  |  |  |  |
| Short | 2.2 | 6.3 | 5.7 | 9.2 | 21.9 | 17.9 | 9.3 | 21.7 | 23.0 | 8.3 | 20.1 | 18.4 | 30.3 | 52.7 | 50.9 | -0,01 |
| Medium | 37.7 | 47.8 | 47.4 | 49.2 | 51.8 | 53.4 | 53.5 | 55.7 | 51.2 | 46.9 | 51.0 | 51.2 | 49.7 | 38.2 | 37.9 | -0,01 |
| Long | 60.1 | 45.8 | 46.9 | 41.7 | 26.3 | 28.6 | 37.2 | 22.6 | 25.8 | 44.8 | 28.9 | 30.4 | 20.0 | 9.1 | 11.2 | 0,02 |
| Preterm |  |  |  |  |  |  |  |  |  |  |  |  |  |  |  |  |
| >37 weeks | 5.5 | 4.7 | 4.5 | 5.2 | 5.7 | 6.0 | 5.1 | 5.5 | 6.3 | 6.4 | 7.0 | 8.1 | 9.7 | 10.8 | 10.1 | -0,00 |
| Psychiatric diagnosis before age of 18 |  |  |  |  |  |  |  |  |  |  |  |  |  |  |  |  |
| Yes | 4.8 | 4..7 | 4.5 | 5.8 | 5.6 | 5.7 | 6.3 | 5.8 | 5.1 | 9.0 | 8.7 | 8.9 | 15.9 | 14.1 | 12.2 | 0,00 |

^a^ U represents the unweighted sample
^b^ W represents the weighted sample
^c^ B represents the background population.

Note: We also checked for balance when including interaction terms, which also indicated that the covariates were balanced.

1.	Stuart EA, Lee BK, Leacy FP. Prognostic score–based balance measures for propensity score methods in comparative effectiveness research. J Clin Epidemiol. 2013 Aug;66(8 0):S84-S90.e1.

Table 4B Weighted and unweighted estimates by childhood adversity trajectory groups.

|  | DNBC-18 | | | | | | | | | | | | | | | |
| --- | --- | --- | --- | --- | --- | --- | --- | --- | --- | --- | --- | --- | --- | --- | --- | --- |
|  | Frequent Binge drinking | | | | Cannabis use | | | | Drug use | | | | Unsafe sex | | | |
| Childhood adversity Group | OR  (95% CI) – weighted | Percentage points difference (95% CI) – weighted | OR  (95% CI) – Unweighted | Percentage points difference (95% CI) – Unweighted | OR  (95% CI) – weighted | Percentage points difference (95% CI) – weighted | OR  (95% CI) – Unweighted | Percentage points difference (95% CI) – Unweighted | OR  (95% CI) – weighted | Percentage points difference (95% CI) – weighted | OR  (95% CI) – Unweighted | Percentage points difference -Unweighted | OR  (95% CI) – weighted | Percentage points difference (95% CI) – weighted | OR  (95% CI) – Unweighted | Percentage points difference (95% CI)– Unweighted |
| Low adversity | Ref | Ref | Ref | Ref | Ref | Ref | Ref | Ref | Ref | Ref | Ref | Ref | Ref | Ref | Ref | Ref |
| Early life MD | 0.87 (0.78-0.97) | -1.8  (-3.1;  -0.5) | 0.97 (0.89-1.04) | -0.7  (-2.2-0.9) | 1.22 (1.11-1.33) | 3.6  (1.9-  5.4) | 1.28 (1.18-1.38) | 4.46 (2.93-5.99) | 1.18  (1.02-  1.36) | 1.3  (0.1-  2.5) | 1.14 (1.01-1.29) | 0.95 (0.00-1.89) | 1.16 (1.07-1.25) | 3.5  (1.6  -5.5) | 1.11 (1.03-1.19) | 2.5  (0.8- 4.2) |
| Persistent MD | 0.84 (0.70-0.99) | -2.3 (-4.4;  -0.2) | 1.02 (0.90-1.15) | 0.4  (-2.0-2.8) | 1.21 (1.05-1.39) | 3.5  (0.8-  6.2) | 1.27 (1.13-1.44) | 4.45 (2.05-6.85) | 1.26  (1.02-1.57) | 1.9  (0.0-  3.8) | 1.22 (1.00-1.47) | 1.46  (-0.06-2.98) | 1.17 (1.04-1.33) | 3.9  (0.9-  6.9) | 1.17 (1.05-1.30) | 3.7  (1.1- 6.4) |
| Loss or threat of loss | 0.87 (0.77-0.98) | -1.9  (-3.4;  -0.4) | 1.00 (0.92-1.09) | 0.1  (-1.6-1.8) | 1.46 (1.33-1.61) | 7.3  (5.3-  9.3) | 1.44 (1.32-1.57) | 6.90 (5.20-8.61) | 1.76  (1.53-  2.02) | 5.2  (3.7-  6.7) | 1.62 (1.43-1.83) | 3.99 (2.82-5.16) | 1.42 (1.30-1.55) | 8.7  (6.5-  10.8) | 1.38 (1.28-1.49) | 7.8  (6.0- 9.7) |
| High adversity | 0.57 (0.37-0.87) | -6.3  (-10.2;  -2.4) | 0.73 (0.55-0.96) | -5.9  (-10.5- (-)1.3) | 1.59  (1.21-2.09) | 9.1  (3.2-  15.0) | 1.51 (1.17-1.92) | 7.81 (2.66-12.95) | 2.44  (1.71-  3.49) | 9.4  (4.4-14.3) | 1.80 (1.28-2.48) | 5.08 (1.60-8.55) | 1.72 (1.34-2.22) | 13.4  (7.2-  19.7) | 1.51 (1.20-1.89) | 10.0 (4.5-15.6) |

Note. Adjusted for sex, highest household education, maternal age at birth, and preterm birth.

### **Supplementary Table 5. Substance admission separated by alcohol and drug admissions.**

|  | Alcohol admissions | | | Drug admissions | | |
| --- | --- | --- | --- | --- | --- | --- |
| Childhood adversity Group | HR  (95% CI) | HD per 10,000 (95% CI) | Cases | HR  (95% CI) | HD per 10,000 (95% CI) | Cases |
| Low adversity | Ref | Ref | 10822 | Ref | Ref | 2819 |
| Early life MD | 1.25 (1.21-1.29) | 6  (5-7) | 5780 | 1.46 (1.38-1.55) | 3  (2-3) | 1908 |
| Persistent MD | 1.46 (1.41-1.51) | 12  (11-13) | 4637 | 1.76 (1.65-1.88) | 5  (4-6) | 1672 |
| Loss or threat of loss | 1.65 (1.58-1.71) | 17  (15-18) | 3190 | 2.52 (2.36-2.69) | 11  (10-12) | 1350 |
| High adversity | 2.73 (2.60-2.86) | 51  (48-55) | 2285 | 5.38 (5.01-5.77) | 38  (36-41) | 1349 |

Note. Adjusted for sex, highest household education, parental origin, maternal age at birth, and preterm birth.

### **Supplementary Table 6A-B. Sensitivity analysis. Main results after excluding parental education as a control variable.**

Table 7A. HRs and HDs per 10,000 for substance-related ER-admissions, excluding parental education as a control variable.

|  | Registry outcomes | | | | | |
| --- | --- | --- | --- | --- | --- | --- |
|  | Substances | | Violence | | Unintentional Injury | |
| Childhood adversity Group | HR  (95% CI) | HD per 10,000 (95% CI) | HR  (95% CI) | HD per 10,000 (95% CI) | HR  (95% CI) | HD per 10,000 (95% CI) |
| Low adversity | Ref | Ref | Ref | Ref | Ref | Ref |
| Early life MD | 1.39  (1.34-1.44) | 12  (11-13) | 1.46  (1.43-1.49) | 34  (32-36) | 1.11  (1.10-1.12) | 145  (138-152) |
| Persistent MD | 1.70  (1.63-1.76) | 22  (20-23) | 1.93  (1.89-1.98) | 71  (68-73) | 1.20  (1.19-1.21) | 275  (267-283) |
| Loss or threat of loss | 1.95  (1.87-2.03) | 30  (28-31) | 1.84  (1.79-1.89) | 62  (59-65) | 1.24  (1.22-1.25) | 321  (311-331) |
| High adversity | 3.94  (3.74-4.15) | 95  (91-100) | 3.35  (3.24-3.46) | 189  (183-195) | 1.49  (1.47-1.52) | 711  (693-729) |

Note. Adjusted for sex, parental origin, maternal age at birth, and preterm birth

Table 6B. ORs and percentage points difference per 10,000 for substance-related ER-admissions, excluding parental education as a control variable.

|  | DNBC-18 outcomes (weighted) | | | | | | | |
| --- | --- | --- | --- | --- | --- | --- | --- | --- |
|  | Frequent binge drinking | | Cannabis use | | Drug use | | Unsafe sex | |
| Childhood adversity Group | OR  (95% CI) | Percentage points difference (95% CI) | OR  (95% CI) | Percentage points difference (95% CI) | OR  (95% CI) | Percentage points difference (95% CI) | OR  (95% CI) | Percentage points difference (95% CI) |
| Low adversity | Ref | Ref | Ref | Ref | Ref | Ref | Ref | Ref |
| Early life MD | 0.87 (0.78–0.97) | -1.80  (-3.1–  (-)0.5) | 1.22  (1.11–  1.33) | 3.6  (1.9–  5.4) | 1.18  (1.02–  1.36) | 1.3  (0.1–  2.5) | 1.16  (1.07–  1.25) | 3.5  (1.6–  5.5) |
| Persistent MD | 0.84 (0.70–0.99) | -2.3  (-4.4–  (-)0.2) | 1.21  (1.05–  1.39) | 3.5  (0.8–  6.2) | 1.26  (1.02–  1.57) | 1.9  (0.0–  3.8) | 1.17  (1.04–  1.33) | 3.9  (0.9–  6.9) |
| Loss or threat of loss | 0.87 (0.77–0.98) | -1.9  (-3.4–  (-)0.4 | 1.46  (1.33–  1.61) | 7.3  (5.3– 9.3) | 1.76  (1.53–  2.02) | 5.2  (3.7–  6.7) | 1.42  (1.30–  1.55) | 8.7  (6.5–  10.8) |
| High adversity | 0.57 (0.37–0.87) | -6.3  (-10.2–  (-)2.4) | 1.59  (1.21–  2.09) | 9.1  (3.2–  15.0) | 2.44  (1.71–  3.49) | 9.4  (4.4–  14.3) | 1.72  (1.34–  2.22) | 13.4  (7.2–  19.7) |

Note. Adjusted for sex, maternal age at birth, and preterm birth

### **Supplementary Table 7. Sensitivity analysis. Comparison of incidence rate ratio (IRR) for violence and unintentional injury admissions in the background population, weighted sample, and unweighted sample. The estimates are not adjusted for covariates.**

|  | Low adversity | | Early life MD | | Persistence MD | | Loss or threat of loss | | High adversity | |
| --- | --- | --- | --- | --- | --- | --- | --- | --- | --- | --- |
|  | IRR (95% CI) | | IRR (95% CI) | | IRR (95% CI) | | IRR (95% CI) | | IRR (95% CI) | |
|  | Violence | Unintentional Injury | Violence | Unintentional Injury | Violence | Unintentional Injury | Violence | Unintentional Injury | Violence | Unintentional Injury |
| Unweighted | Ref | Ref | 0.97  ( 0.58- 1.62) | 1.02  (0.94- 1.11) | 1.80  (0.98- 3.32) | 0.98  (0.87- 1.11) | 2.03  (1.36- 3.04) | 1.36  (1.24- 1.49) | 3.29  (1.35- 7.99) | 1.47  (1.15- 1.88) |
| Cases/PY | 127/ 72216 | 6980/ 72216 | 18/ 10602 | 1049/ 10602 | 13/ 4061 | 385/ 4061 | 31/ 8656 | 1135/ 8656 | 5/ 878 | 125/ 878 |
| Weighted | Ref | Ref | 0.89  (0.51-  1.55) | 1.09  (0.99-  1.19) | 2.01  (1.05-  3.85) | 1.00  (0.87-  1.14) | 2.37  (1.49-  3.77) | 1.40  (1.27-  1.54) | 2.95  (1.08-  8.05) | 1.67  (1.27-  2.21) |
| Cases/PY | 141/  68872 | 7027/  68872 | 22/  12193 | 1353/  12193 | 20/  4723 | 480/  4723 | 47/  9726 | 1385/  9726 | 9/  1447 | 248/  1447 |
| Background | Ref | Ref | 1.65  (1.55-  1.76) | 1.10  (1.09-  1.11) | 2.12  (1.97-  2.29) | 1.10  (1.08-  1.12) | 2.18  (2.04-  2.33) | 1.33  (1.31-  1.35) | 4.51  (4.14-  4.91) | 1.68  (1.65-  1.72) |
| Cases/PY | 3116/  846528 | 99281/  846528 | 1302/  213475 | 27465/  213475 | 883/  112668 | 14542/  112668 | 1229/  153229 | 23919/  153229 | 638/  38355 | 755/  38355 |

### **Supplementary Table 8A-E. Results of main analysis stratified by sex.**

Table 8A. HRs and HDs 10,000 for substance ER-admissions stratified sex.

|  | Substance use | | | | | |
| --- | --- | --- | --- | --- | --- | --- |
| Childhood adversity Group | HR  (95% CI) – Male | HD per 10,000 (95% CI) – Male | Cases/  Person year -  Male | HR  (95% CI) – Female | HD per 10,000 (95% CI) – Female | Cases/  Person year -  Female |
| Low adversity | Ref | Ref | 7270 / 2236947 | Ref | Ref | 6157 / 2099801 |
| Early life MD | 1.27 (1.22–1.33) | 8.7 (7.08-10.3) | 4029 /  889460 | 1.31 (1.24–1.38) | 8.98 (7.39-10.6) | 3508/ 840770 |
| Persistent MD | 1.50 (1.43–1.58) | 16.9 (14.8-19.) | 3325 /  595312 | 1.53 (1.44–1.62) | 16. (13.9-18.1) | 2853/ 563579 |
| Loss or threat of loss | 1.77 (1.67–1.87) | 26.1 (23.5-28.7) | 2319 /  372625 | 1.88 (1.76–2.00) | 26.9 (24.3-29.5) | 2119/ 356102 |
| High adversity | 3.09 (2.88–3.32) | 83 (77.3-89.1) | 1897 /  148033 | 3.51 (3.22–3.83) | 90.2 (83.8-96.6) | 1626/ 124172 |

Note. Adjusted for sex, highest household education, parental origin, maternal age at birth, and preterm birth.

Table 8B. HRs and HDs per 10,000 for violence ER-admissions stratified by sex.

|  | Violence | | | | | |
| --- | --- | --- | --- | --- | --- | --- |
| Childhood adversity Group | HR  (95% CI) – Male | HD per 10,000 (95% CI) – Male | Cases/  Person year -  Male | HR  (95% CI) – Female | HD per 10,000 (95% CI) – Female | Cases/  Person year -  Female |
| Low adversity | Ref | Ref | 2642 /  2236947 | Ref | Ref | 5882/ 2099801 |
| Early life MD | 1.27 (1.24–1.30) | 31 (28–34) | 14872 /  889460 | 1.61 (1.54–1.69) | 17 (15–19) | 4642/ 840770 |
| Persistent MD | 1.56 (1.52–1.60) | 72 (68–76) | 13106 /  595312 | 2.17 (2.06–2.28) | 37 (35–40) | 4617/ 563579 |
| Loss or threat of loss | 1.51 (1.46–1.55) | 62 (57–66) | 7199 / 372625 | 2.44 (2.31–2.57) | 43.9 (41–47) | 2858/ 356102 |
| High adversity | 2.23 (2.14–2.32) | 177 (167–187) | 5059 / 148033 | 4.60 (4.31–4.92) | 145 (137–153) | 2481/ 124172 |

Note. Adjusted for sex, highest household education, parental origin, maternal age at birth, and preterm birth.

Table 8C. HRs and HDs per 10,000 for unintentional injury ER-admissions stratified by sex.

|  | Unintentional injuries | | | | | |
| --- | --- | --- | --- | --- | --- | --- |
| Childhood adversity Group | HR (95% CI) – male | HD per 10,000 (95% CI) – male | Cases/  Person year -  Male | HR (95% CI) – female | HD per 10,000 (95% CI) – female | Cases/  Person year -  Female |
| Low adversity | Ref | Ref | 355611 / 2236947 | Ref | Ref | 232607 / 2099801 |
| Early life MD | 1.05 (1.04–1.06) | 74 (63-84) | 160537 / 889460 | 1.04 (1.03–1.05) | 44 (35–53) | 103696 / 840770 |
| Persistent MD | 1.12 (1.11–1.14) | 211 (198–224) | 119023 / 595312 | 1.07 (1.05–1.08) | 79 (68–90) | 72746 / 563579 |
| Loss or threat of loss | 1.14 (1.13–1.15) | 226 (211–241) | 71491 / 372625 | 1.15 (1.13–1.17) | 266 (253–279) | 51240 / 356102 |
| High adversity | 1.25 (1.22–1.27) | 451 (425–477) | 35041 / 148033 | 1.41 (1.37–1.46) | 526 (501–551) | 23043 / 124172 |

Note. Adjusted for sex, highest household education, parental origin, maternal age at birth, and preterm birth.

Table 8D. ORs and percentage points difference for binge drinking, drug use, and unsafe sex stratified by sex

| DNBC-18 (weighted) | | | | | | | | | | | | | | | | |
| --- | --- | --- | --- | --- | --- | --- | --- | --- | --- | --- | --- | --- | --- | --- | --- | --- |
|  | Frequent Binge drinking | | | | Cannabis use | | | | Drug use | | | | Unsafe sex | | | |
| Childhood adversity Group | OR  (95% CI) – Male | Percentage points difference (95% CI)  – Male | OR (95% CI) – Female | Percentage points difference (95% CI) – Female | OR  (95% CI) – Male | Percentage points difference (95% CI)  – Male | OR (95% CI) – Female | Percentage points difference (95% CI) – Female | OR  (95% CI) – Male | Percentage points difference (95% CI) – Male | OR  (95% CI) – Female | Percentage points difference (95% CI) – Female | OR  (95% CI) – Male | Percentage points difference (95% CI) – Male | OR  (95% CI) – Female | Percentage points difference (95% CI) – Female |
| Low adversity | Ref | Ref | Ref | Ref | Ref | Ref | Ref | Ref | Ref | Ref | Ref | Ref | Ref | Ref | Ref | Ref |
| Early life MD | 0.88  (0.76-  1.02) | -2.1  (-4.3-  0.2) | 1.07  (0.92-  1.25) | 0.7  (-0.9-  2.4) | 1.11 (0.98-  1.26) | 2.2  (-0.5-  4.9) | 1.44  (1.28-  1.63) | 5.9  (3.8-  8.0) | 0.98  (0.81-  1.19) | -0.2  (-2.1-  1.7) | 1.38  (1.12-  1.71) | 1.8  (0.5-  3.0) | 1.02  (0.90-  1.15) | 0.4  (-2.5-  3.3) | 1.18  (1.07-  1.31) | 4.0  (1.5-  6.5) |
| Persistent MD | 0.93  (0.74-  1.18) | -1.1  (-4.8-  2.6) | 0.90  (0.70-  1.15) | -1.1  (-3.4-  1.3) | 1.12  (0.92-  1.38) | 2.5  (-1.9-  6.8) | 1.40  (1.16-  1.69) | 5.4  (2.1-  8.7) | 1.03  (0.76-  1.38) | 0.3  (-2.7-  3.3) | 1.50  (1.10-  2.05) | 2.3  (0.2-  4.3) | 0.98  (0.81-  1.18) | -0.5  (-5.1-  4.1) | 1.23  (1.05-  1.44) | 5.0  (1.2-  8.9) |
| Loss or threat of loss | 0.90  (0.76-  1.06) | -1.7  (-4.3-  0.9) | 1.00  (0.85-  1.18) | 0.0  (-1.7-  1.7) | 1.41  (1.23-  1.62) | 7.6  (4.4-  10.8) | 1.59  (1.40-  1.81) | 7.7  (5.3-  10.0) | 1.55  (1.29-  1.86) | 5.1  (2.7-  7.6) | 1.89  (1.54-  2.33) | 4.0  (2.4-  5.6) | 1.30  (1.14-  1.49) | 6.5  (3.2-  9.8) | 1.41  (1.26-  1.57) | 8.3  (5.6-  11.0) |
| High adversity | 0.43  (0.22-  0,84) | -10.6  (-16.7-  -4.4) | 1.26  (0.74-  2.14) | 2.6  (-3.9-  9.0) | 1.45  (0.98-  2.15) | 8.3  (0.9-17.4) | 1.89  (1.29-  2.77) | 11.1  (3.4-  18.7) | 1.75  (1.07-  2.86) | 6.9  (0.3-  14.2) | 2.73  (1.64-4.53) | 7.5  (2.1-  12.8) | 1.22  (0.84-  1.78) | 4.9  (-4.4-  14.2) | 1.96  (1.39-  2.76) | 16.5  (8.3-  24.8) |

Note. Adjusted for sex, highest household education, maternal age at birth, and preterm birth.

Table 8E. Number of affirmative answers and proportions by adversity groups for frequent binge drinking, drug use, and unsafe sex (weighted).

| DNBC-18 (weighted) | | | | | | | | |
| --- | --- | --- | --- | --- | --- | --- | --- | --- |
|  | Frequent binge drinking | | Cannabis use | | Drug use | | Unsafe sex | |
| Childhood adversity Group | Male | Female | Male | Female | Male | Female | Male | Female |
| Low adversity | 2609 (21.1) | 1419 (12.0) | 3654 (29.6) | 2061 (17.4) | 1379 (11.2) | 566 (4.8) | 5111 (41.4) | 4769 (40.3) |
| Early life MD | 386 (17.8) | 251 (11.7) | 674 (31.0) | 505 (23.5) | 248 (11.4) | 155 (7.2) | 939 (43.2) | 1009 (46.9) |
| Persistent MD | 150 (18.5) | 84 (9.9) | 250 (30.9) | 196 (23.2) | 95 (11.8) | 67 (7.9) | 343 (42.4) | 408 (48.3) |
| Loss or threat of loss | 297 (18.3) | 195 (11.0) | 593 (36.5) | 446 (25.3) | 273 (16.8) | 168 (9.6) | 796 (49.0) | 894 (50.7) |
| High adversity | 24 (8.6) | 30 (11.9) | 98 (35.9) | 75 (30.0) | 54 (19.5) | 39 (15.7) | 136 (49.6) | 157 (62.9) |
